# Association of E484K and L452R spike protein mutations with SARS-CoV-2 infection in vaccinated persons---Maryland, January – May 2021

**DOI:** 10.1101/2021.07.29.21261006

**Authors:** Kenneth A. Feder, Ami Patel, Venkata R. Vepachedu, Catherine Dominguez, Eric N. Keller, Liore Klein, Curi Kim, Tim Blood, Judie Hyun, Theo Williams, Katherine A. Feldman, Heba H. Mostafa, C. Paul Morris, Jacques Ravel, Monique Duwell, David Blythe, Robert Myers

## Abstract

**Background:** The E484K and L452R amino acid substitutions on the spike protein of SARS-CoV-2 are associated with reduced neutralization by antibodies from acquired immunity. This study examines the respective association of these mutations with infection in persons who had previously received a COVID-19 vaccine.

**Methods:** Genetic sequences from SARS-CoV-2 specimens collected from Maryland residents and reported to Maryland Department of Health were linked to vaccination history. The prevalence of infections in fully vaccinated persons -- defined as being at least two weeks past receiving the final scheduled dose of a COVID-19 vaccine series -- was compared between infections caused by viruses carrying E484K to those not carrying E484K, and between infections caused by viruses carrying L452R to those not carrying L452R, using logistic regression to adjust for confounding.

**Results:** Of 9,048 sequenced SARS-CoV-2 specimens examined, 265 (2.9%) were collected from fully vaccinated persons. In adjusted analysis, the E484K substitution was associated with an increase in the odds of the sequenced specimen being collected from a fully vaccinated person (OR 1.96, 95% CI, 1.36 to 2.83). The L452R mutation was not significantly associated with infections in vaccinated persons (OR 1.07, 95% CI, 0.69 to 1.68).

**Conclusion:** Though more than 97% of SARS-CoV-2 infections were in persons who were not fully vaccinated, the E484K mutation was associated with increased odds of SARS-CoV-2 infection in vaccinated persons. Linking vaccination and sequencing data can help identify and estimate the impact SARS-CoV-2 mutations may have on vaccine effectiveness.

**Summary:** In viruses sequenced for Maryland’s routine SARS-CoV-2 genomic surveillance, the spike protein amino acid substitution E484K was more prevalent in viruses that infected vaccinated people than in viruses that infected people who were not vaccinated.

## 1. Introduction

With the advent of safe and efficacious COVID-19 vaccines (1-5) comes concern that mutations in the SARS-CoV-2 virus will erode the effectiveness of these vaccines. The United States Centers for Disease Control and Prevention (CDC U.S. Government SARS-CoV-2 Interagency Group) has highlighted two spike protein mutations that are among those concerning for their possible impact on protection from acquired immunity: substitution of lysine for glutamic acid at the 484 position (E484K); and substitution of arginine for leucine at the 452 position (L452R) (6).

E484K has emerged independently in multiple virus lineages, including several lineages designated by CDC as variants of concern or interest such as B.1.351 (labeled “Beta” variant by WHO), P.1 (“Gamma”), B.1.525 (“Eta”), and B.1.526 (“Iota”) (6). The B.1.351 lineage has been associated with reduced neutralization by monoclonal antibodies, convalescent plasma, and sera from vaccinated persons in laboratory studies (7-10). In South Africa – where B.1.351 was the dominant strain – multiple SARS-CoV-2 vaccines showed reduced effectiveness as compared to clinical trials in other countries (3,4). Similarly, the P.1 lineage is less likely to be neutralized by convalescent plasma in laboratory studies (11). Some SARS-CoV-2 vaccine trials conducted in Brazil, where P.1 was a common lineage, demonstrated reduced effectiveness as compared to trials in other countries (4). By contrast, an observational study on the association of B.1.526 lineage with infections in vaccinated persons did not show any association; however, relatively few specimens were sequenced over the study period (12).

L452R has also emerged independently in multiple SARS-CoV-2 lineages, including several designated by CDC to be variants of interest or concern such as B.1.427/B.1.429 (“Epsilon”), B.1.526.1, B.1.617.1 (“Kappa”), and B.1.617.2 (“Delta”), as well as other lineages detected in the United States such as B.1, A.2.5, and C.37 (“Lambda”). In particular, B.1.427, B.1.429, and B.1.617.1 were associated with reduced antibody neutralization with sera from vaccinated persons (13,14), and B.1.617.2 was associated with reduced neutralization with sera from persons vaccinated with BNT162b2 with an effect size comparable degree to B.1.351 (10).

To date, no study has directly examined the association of these two mutations with infection in vaccinated persons in the general population combining evidence across multiple lineages and vaccine types. One study, from Israel, found increased rates of infection with the B.1.351 variant specifically in persons vaccinated with the BNT162b2 mRNA vaccine (15).

This study examines the association of the E484K and L452R substitutions with infections in vaccinated persons using SARS-CoV-2 genetic surveillance data from the U.S. State of Maryland. This study examines the following questions: 1) What were the prevalences of the E484K and L452R substitutions in Maryland in 2021, and which SARS-CoV-2 lineages primarily carried those substitutions? 2) Were persons infected with viruses carrying the E484K or L452R substitutions more likely to have been vaccinated prior to infection, as compared to persons infected with viruses not carrying the E484K or L452R substitutions? 3) Did any observed association of the E484K or L452R with post-vaccination infection remain if only infections that were followed by hospitalization (i.e. severe or critical infections) are counted? 4) Did any observed association of E484K or L452R with post-vaccination infection differ by vaccine type?

## 2. Methods

### 2.1 Data

This retrospective analysis uses all specimens on GISAID collected between the start of January 2021 and the second week of May of 2021 that were a) listed as being collected from Maryland residents and b) could be successfully linked to a case record in Maryland Department of Health’s COVID-19 surveillance systems using identifying information reported by the sequencing labs to the Department. These 9,048 specimens represent 89% of Maryland resident specimens published on GISAID during that time period, and 5.3% of all Maryland’s SARS-CoV-2 infections confirmed by nucleic acid amplification test (NAAT) during that time period. Note that each specimen was collected from a unique patient, so the unit of analysis is patients.

### 2.2. Measures

#### 2.2.1. Outcomes

##### 2.2.1.1. Infection with or without hospitalization in persons who were fully vaccinated

Infection in a person who was fully vaccinated (n = 265) was defined as having a SARS-CoV-2 respiratory specimen positive by NAAT or antigen test collected 14 or more days after receiving the final scheduled dose of a COVID-19 vaccine series. For the BNT162b2 mRNA COVID-19 vaccine and the mRNA-1273 COVID-19 vaccine this meant 14 or more days following receipt of a second dose; for the Ad26.COV2.S COVID-19 vaccine this meant 14 or more days following receipt of the single dose. Hospitalization of a fully vaccinated person (n=35) was defined the same, with the added condition that the patient was admitted to the hospital within 28 days following collection of their first SARS-CoV-2 positive specimen.

##### 2.2.1.2. Infection with or without hospitalization in persons who received any vaccination

Infection in persons with any vaccination (n = 554) was defined as having SARS-CoV-2 positive respiratory specimen collected 14 or more days after receiving a first dose of any COVID-19 vaccine. Note that this includes all instances of infection in fully vaccinated persons, plus any additional instances of infection that occurred fourteen or more days following inoculation with a single dose of BNT162b2 or mRNA-1273 but prior to completion of either of those vaccine series. Hospitalization of a person with any vaccination (n =105) was defined the same, with the added condition that the patient was admitted to the hospital within 28 days following collection of their first SARS-CoV-2 positive specimen.

#### 2.2.2. Primary comparison groups

Persons who were infected with SARS-CoV-2 viruses that carried the E484K substitution were compared to persons who were infected with SARS-CoV-2 viruses that did not carry the E484K substitution. Persons who were infected with SARS-CoV-2 viruses carrying the L452R substitution were compared to persons infected with SARS-CoV-2 viruses not carrying the L452R substitution. (There were no viruses that carried both substitutions.)

#### 2.2.3. Other covariates

To adjust for confounding, the following covariates were examined and included in adjusted analyses: age group, gender, region of residence, laboratory submitting to GISAID, and week of specimen collection.

### 2.3 Analytic Approach

#### 2.3.1 Descriptive Analysis

We plotted the prevalence of viruses carrying the E484K and L452R substitutions by week of specimen collection, stratified by the virus PANGO Lineage (version 3.0.2) listed on GISAID. We also calculated the prevalence of all study outcomes and covariates in the whole sample, and in subsamples stratified by the presence or absence of each substitution of interest.

#### 2.3.2 Post-Vaccination Infection

Using logistic regression, we then estimated the respective crude and adjusted associations of each of the two substitutions with each “infection in a vaccinated person” outcome (see 2.2) using logistic regression. In the adjusted logistic regression, we included dummy variables to adjust for potential confounding covariates (see section 2.2.3). Regression coefficients were exponentiated, and interpreted as odds ratios. Wald confidence intervals (95%) were estimated for all regression coefficients; odds ratios with 95% confidence intervals not containing 1 were considered statistically significant at the p<0.05 level.

#### 2.3.3 Post-Vaccination Hospitalization

The analysis described in 2.3.2 was repeated, but the outcome was restricted to cases of post-vaccination infection that were followed by hospitalization within 28 days of specimen collection.

#### 2.3.4 Vaccine-Specific Sub-Analysis

The analysis described in 2.3.2 was repeated, but the outcome was restricted to infection in a vaccinated person who was vaccinated with a specific vaccine. This analysis was done for all three vaccine types. For each analysis, persons vaccinated with a vaccine other than the type examined in that analysis were excluded (e.g., persons vaccinated with mRNA-1273 or Ad26.COV2.S vaccines were excluded from analysis of the BNT162b2 vaccine). Note that, because Ad26.COV2.S requires only one dose, by definition, the only outcome that could be examined was infection in a fully vaccinated person.

#### 2.3.5 Sensitivity Analysis

First, Maryland’s public health laboratory actively tries to obtain SARS-CoV-2 specimens for sequencing from persons who were vaccinated prior to their infection, so the proportion of post-vaccination infections in specimens sequenced at that laboratory exceeds what would be expected in other laboratories. In the first sensitivity analysis, we repeat the main analysis excluding all specimens sequenced by Maryland’s public health laboratory.

Second, among the most important potential confounders in this analysis is date of collection, because amino acid substitutions that became more prevalent later in the study period might appear to be associated with vaccination simply because more people had been vaccinated by that time. In the second sensitivity analyses, we instead adjusted for time using a cubic polynomial basis, rather than using dummy variables for week of collection, to ensure findings are robust to how we chose to model time.

Third, some of the assumptions of Wald confidence intervals for logistic regression coefficients may be violated for analyses where the outcome is very rare. For the third sensitivity analysis, the analysis was repeated using ordinary bootstrapping, with 1,000 bootstrap replicates.

#### 2.3.6 Software

All analyses were conducted in R version 3.6.1.

### 2.4 Research Determination

Case investigation, data collection, and analysis were conducted for public health purposes. This project was reviewed by the Division of Scientific Education and Professional Development within the Center for Surveillance, Epidemiology, and Laboratory Services at the Centers for Disease Control and Prevention (CDC). The project was determined to meet the requirements of public health surveillance covered by the U.S. Department of Health and Human Services Policy for the Protection of Human Research Subjects as defined in 45 C.F.R. part 46.102(l)(2), 21 C.F.R. part 56; 42 U.S.C. Sec. 241(d); 5 U.S.C. Sec. 552a; 44 U.S.C. Sec. 3501 et seq. In., and the decision was made that this project was nonresearch and did not require ethical review by the CDC Human Research Protection Office. Ethical approval was waived and informed consent was not required.

## Results

### 3.1 Descriptive Analysis

#### 3.1.1 Amino Acid Substitutions of Interest

Out of 9,048 SARS-CoV-2 specimens on GISAID collected from Maryland residents during the study period and included in this analysis, 1,187 (13.1%) carried the E484K substitution and 731 (8.1%) carried the L452R substitution (Table 1). The most common lineages carrying E484K were B.1.526 (33.3% of specimens carrying E484K), R.1 (27.0%) and B.1.351 (9.7%). The most common lineages carrying L452R were B.1.526.1 (64.6%), B.1.429 (15.2%) and B.1 (6.2%). (Figure 1)

**Table 1.** Prevalence SARS-CoV-2 infection in vaccinated persons with and without hospitalization and other demographics in Maryland residents (n=9,048) with sequenced SARS-CoV-2 specimens collected January-May 2021.

|  | All |  | E484K Absent |  | E484K Present |  | L452R Absent |  | L452R Present |  |
| --- | --- | --- | --- | --- | --- | --- | --- | --- | --- | --- |
|  | Count | Percent | Count | Percent | Count | Percent | Count | Percent | Count | Percent |
| Population Totals (Percent of All) | 9,048 |  | 7,861 | 86.9% | 1,187 | 13.1% | 8317 | 91.9% | 731 | 8.1% |
| Infections In Vaccinated Persons |  |  |  |  |  |  |  |  |  |  |
| In fully vaccinated persons | 265 | 2.9% | 217 | 2.8% | 48 | 4.0% | 238 | 2.9% | 27 | 3.7% |
| In persons with any vaccination | 554 | 6.1% | 457 | 5.8% | 97 | 8.2% | 498 | 6.0% | 56 | 7.7% |
| In fully vaccinated persons, hospitalized following infection | 35 | 0.4% | 26 | 0.3% | 9 | 0.8% | 34 | 0.4% | 1 | 0.1% |
| In persons with any vaccination, hospitalized following infection | 105 | 1.2% | 80 | 1.0% | 25 | 2.1% | 96 | 1.2% | 9 | 1.2% |
| Gender |  |  |  |  |  |  |  |  |  |  |
| Female | 4785 | 52.9% | 4195 | 53.4% | 590 | 49.7% | 4405 | 53.0% | 380 | 52.0% |
| Male | 4263 | 47.1% | 3666 | 46.6% | 597 | 50.3% | 3912 | 47.0% | 351 | 48.0% |
| Age (years) |  |  |  |  |  |  |  |  |  |  |
| 0-18 | 1420 | 15.7% | 1274 | 16.2% | 146 | 12.3% | 1315 | 15.8% | 105 | 14.4% |
| 18-64 | 6643 | 73.4% | 5742 | 73.0% | 901 | 75.9% | 6093 | 73.3% | 550 | 75.2% |
| 65+ | 985 | 10.9% | 845 | 10.7% | 140 | 11.8% | 909 | 10.9% | 76 | 10.4% |
| Region |  |  |  |  |  |  |  |  |  |  |
| Baltimore Metro Area | 6351 | 70.2% | 5625 | 71.6% | 726 | 61.2% | 5778 | 69.5% | 573 | 78.4% |
| Eastern Shore | 819 | 9.1% | 709 | 9.0% | 110 | 9.3% | 773 | 9.3% | 46 | 6.3% |
| National Capital Region | 1642 | 18.1% | 1323 | 16.8% | 319 | 26.9% | 1548 | 18.6% | 94 | 12.9% |
| Southern Maryland | 105 | 1.2% | 90 | 1.1% | 15 | 1.3% | 99 | 1.2% | 6 | 0.8% |
| Western Maryland | 131 | 1.4% | 114 | 1.5% | 17 | 1.4% | 119 | 1.4% | 12 | 1.6% |
| Sequencing Laboratory |  |  |  |  |  |  |  |  |  |  |
| CDC Contractor | 3164 | 35.0% | 2788 | 35.5% | 376 | 31.7% | 2944 | 35.4% | 220 | 30.1% |
| Johns Hopkins | 2142 | 23.7% | 1910 | 24.3% | 232 | 19.5% | 1965 | 23.6% | 177 | 24.2% |
| University of Maryland | 2305 | 25.5% | 1892 | 24.1% | 413 | 34.8% | 2098 | 25.2% | 207 | 28.3% |
| Maryland Public Health Laboratory | 1362 | 15.1% | 1205 | 15.3% | 157 | 13.2% | 1237 | 14.9% | 125 | 17.1% |
| Other | 75 | 0.8% | 66 | 0.8% | 9 | 0.8% | 73 | 0.9% | 2 | 0.3% |

**Figure 1.**
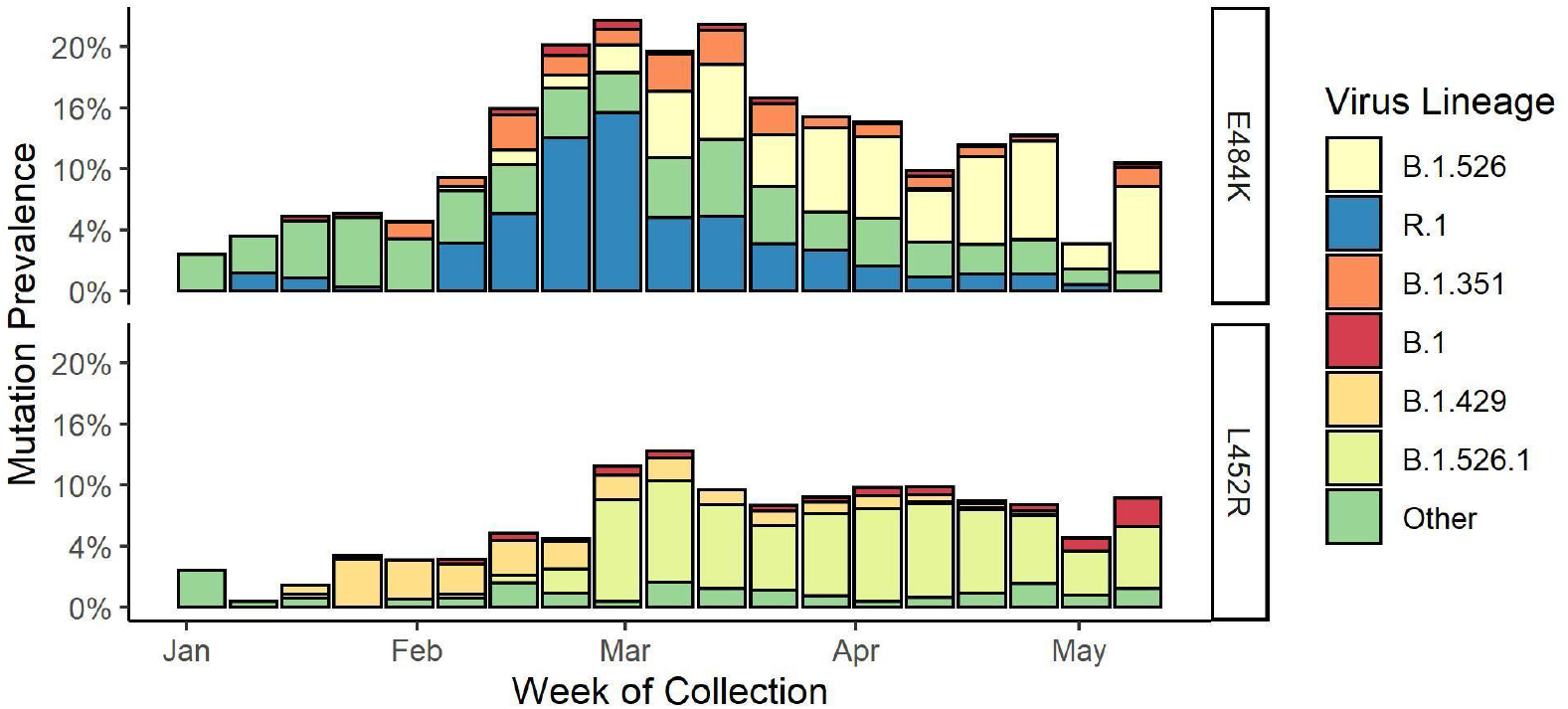
Percent of sequenced SARS-CoV-2 specimens (n=9,048) carrying E484K and L452R mutations by week of collection and lineage, Maryland, January – May 2021

#### 3.1.2 Infections in Vaccinated Persons

Among the 9,048 persons whose sequenced specimens were included in this analysis, there were 265 (2.9%) instances of infection in persons who were fully vaccinated (Table 1): 66.3% were in persons vaccinated with BNT162b2, 25.7% were in persons vaccinated with mRNA-1273, and 7.9% were in persons vaccinated with Ad26.COV2.S COVID-19 (not shown in Table 1). Thirty five (0.4%) infections resulted in hospitalization.

There were 554 (6.1%) instances of infection in persons with any vaccination (including the aforementioned 265 post-full infections). Of these people who had at least one vaccination and were infected, 65.4% were in persons vaccinated with BNT162b2, 30.8% were in persons vaccinated with mRNA-1273, and 3.8% were in persons vaccinated with Ad26.COV2.S COVID-19 (not shown in Table 1). One hundred and five (1.2%) infections resulted in hospitalization (Table 1) (Figure 2).

**Figure 2.**
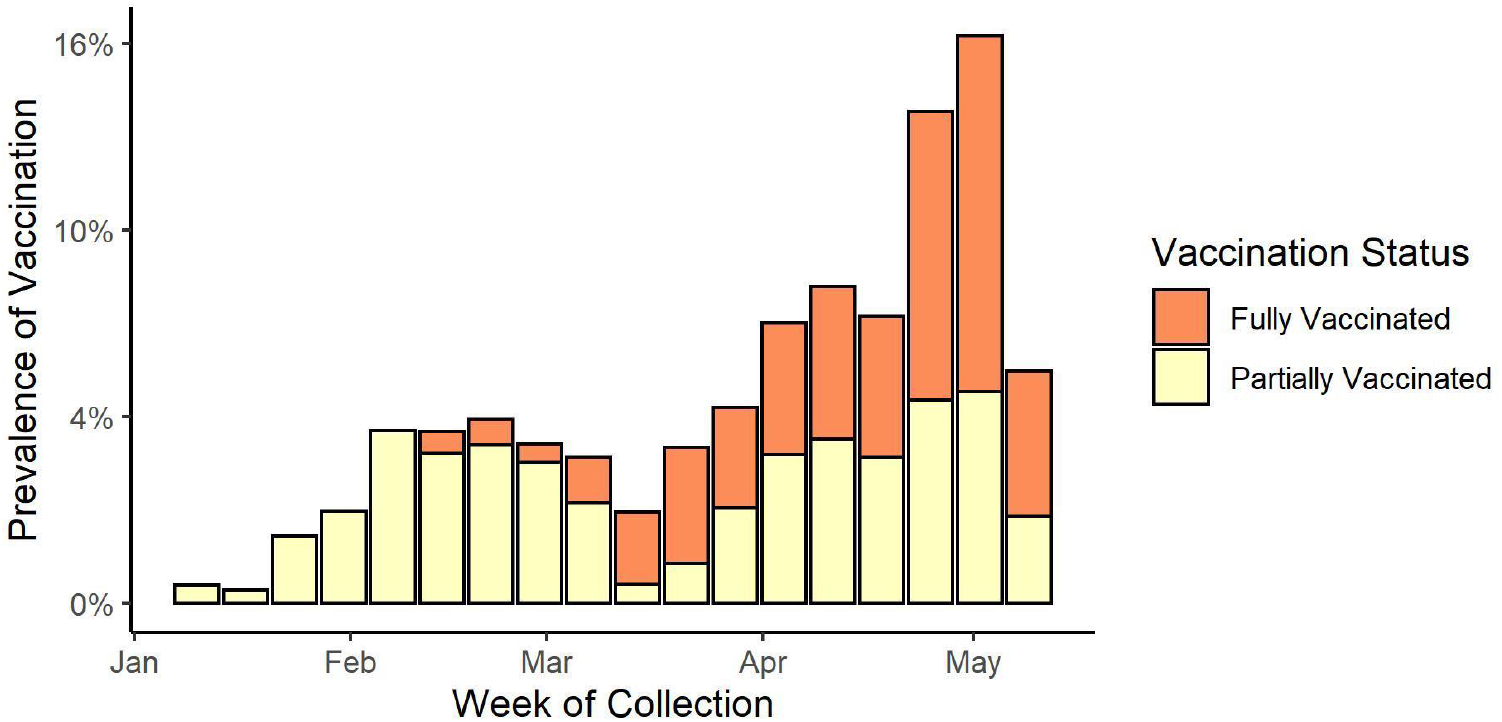
Percent of sequenced SARS-CoV-2 specimens (n=9,048) collected from vaccinated persons by week of collection and vaccination status, Maryland, Jan-May 2021 Fully vaccinated (n=265) means ≥ 14 days past the final scheduled dose of any COVID-19 vaccine series. Partially vaccinated (n=289) means ≥14 days past the first scheduled dose of the BNT162b2 or mRNA-1273, but not yet 14 days past the second scheduled doses of those series.

### 3.2 SARS-CoV-2 Infection in Vaccinated Persons

#### 3.2.1 E484K

Viruses carrying the E484K mutation were significantly more likely to have been collected from fully vaccinated persons (OR 1.48, 95% CI 1.08 to 2.04; aOR 1.96, 95% CI, 1.36 to 2.83) and from persons with any vaccination (OR 1.44, 95% CI, 1.15 to 1.81; aOR 1.68, 95% CI, 1.30 to 2.18) (Table 2).

**Table 2.** Crude and adjusted odds ratios for association of E484K and L452R mutations with post-vaccination SARS-CoV-2 infection with and without hospitalization.

| Association | Odds Ratio (95% CI) |  |
| --- | --- | --- |
|  | Crude | Adjusted |
| **E484K** |  |  |
| In persons with any vaccination | **1.44 (1.15 to 1.81)** | **1.68 (1.3 to 2.18)** |
| In fully vaccinated persons | **1.48 (1.08 to 2.04)** | **1.96 (1.36 to 2.83)** |
| In persons with any vaccination, hospitalized following infection | **2.09 (1.33 to 3.29)** | **2.15 (1.28 to 3.60)** |
| In fully vaccinated persons, hospitalized following infection | **2.3 (1.08 to 4.93)** | **2.56 (1.09 to 6.01)** |
| **L452R** |  |  |
| In persons with any vaccination | 1.3 (0.98 to 1.74) | 1.12 (0.81 to 1.53) |
| In fully vaccinated persons | 1.3 (0.87 to 1.95) | 1.07 (0.69 to 1.68) |
| In persons with any vaccination, hospitalized following infection | 1.07 (0.54 to 2.12) | 0.85 (0.41 to 1.78) |
| In fully vaccinated persons, hospitalized following infection | 0.33 (0.05 to 2.44) | 0.27 (0.04 to 2.06) |

Associations statistically significant at the $p < 0.05$ level are bolded.
Adjusted analyses adjusted for age, sex, region of residence, sequencing laboratory, and week of collection.

#### 3.2.2 L452R

Viruses carrying the L452R mutation were not significantly more likely to be collected from fully vaccinated persons (OR 1.30, 95% CI, 0.87 to 1.95; aOR 1.07, 95% CI, 0.69 to 1.68); or from persons with any vaccination (OR 1.30, 95% CI, 0.98 to 1.74; aOR 1.12, 95% CI, 0.81 to 1.53) (Table 2).

### 3.3 Infection and Hospitalization in Vaccinated Persons

#### 3.3.1 E484K

Viruses carrying the E484K mutation were significantly more likely to have been collected from hospitalized fully vaccinated persons (OR 2.30, 95% CI 1.08 to 4.93; aOR 2.56, 1.09 to 6.01) and hospitalized persons with any vaccination (OR 2.09, 95% CI, 1.33 to 3.29; aOR 2.15, 95% CI 1.28 to 3.60) (Table 2).

#### 3.3.2 L452R

Viruses carrying the L452R mutation were not significantly more likely to be collected from hospitalized fully vaccinated persons (OR 0.33, 95% CI, 0.05 to 2.44; aOR 95% CI, 0.04 to 2.06); or from persons with any vaccination (OR 1.07, 95% CI, 0.54 to 2.12; aOR 0.85, 95% CI, 0.41 to 1.78) (Table 2).

### 3.4 Infection in vaccinated persons vaccinated with a specific vaccine model

In adjusted analysis, for both E484K and L452R, associations with infection in fully vaccinated persons were comparable for all vaccine types (Table 3).

**Table 3.** Crude and adjusted odds ratios for association of E484K and L452R mutations with vaccine-specific post-vaccination SARS-CoV-2 infection.

| Association | Odds Ratio (95% CI) |  |
| --- | --- | --- |
|  | Crude | Adjusted |
| **E484K** |  |  |
| In persons with any vaccination with BNT162b2 | 1.23 (0.92 to 1.64) | **1.47 (1.07 to 2.03)** |
| In persons fully vaccinated with BNT162b2 | 1.38 (0.93 to 2.06) | **1.88 (1.19 to 2.96)** |
| In persons with any vaccination with mRNA-1273 | **1.95 (1.34 to 2.82)** | **2.24 (1.49 to 3.37)** |
| In persons fully vaccinated with mRNA-1273 | 1.78 (0.98 to 3.21) | **2.54 (1.30 to 4.94)** |
| In persons fully vaccinated with Ad26.COV2.S | 1.6 (0.54 to 4.76) | 2.87 (0.89 to 9.26) |
| **L452R** |  |  |
| In persons with any vaccination with BNT162b2 | 1.34 (0.95 to 1.9) | 1.13 (0.78 to 1.64) |
| In persons fully vaccinated with BNT162b2 | 1.3 (0.79 to 2.13) | 1.09 (0.63 to 1.86) |
| In persons with any vaccination with mRNA-1273 | 1.32 (0.8 to 2.20) | 1.22 (0.71 to 2.09) |
| In persons fully vaccinated with mRNA-1273 | 1.57 (0.75 to 3.29) | 1.36 (0.61 to 3.03) |
| In persons fully vaccinated with Ad26.COV2.S | 0.58 (0.08 to 4.32) | 0.61 (0.08 to 4.63) |

Associations statistically significant at the $p < 0.05$ level are bolded.
Adjusted analyses adjusted for age, sex, region of residence, sequencing laboratory, and week of collection.

### 3.5 Sensitivity analysis

Associations estimated in sensitivity analysis did not differ substantially from those in the main analysis (results not shown).

## 4. Discussion

In Maryland, in the first half of 2021, infections in vaccinated persons who received some or all scheduled doses of a COVID-19 vaccine were uncommon. However, when they did occur, these infections in vaccinated persons were disproportionately likely to be infections with viruses that carried the E484K substitution. Consistent with past research (7-11,15,16), this suggests that the three COVID-19 vaccines examined here may have been less effective against viruses carrying the E484K mutation than they were against the population of viruses not carrying this mutation that were circulating in Maryland during early 2021.

By contrast, we did not find an association of L452R with infections in vaccinated persons. As noted, this is in contrast to existing *in vitro* studies on several SARS-CoV-2 lineages that frequently carry L452R; those studies have found lineages carrying L452R were associated with reduced neutralizing titers in convalescent patients and vaccine recipients (13,14). Those studies examined lineages, rather than specific mutations. If L452R was one of several mutations in those lineages contributing to reduced neutralization, its individual contribution might be too small to detect in this study. In short, while the results here are encouraging, they do not obviate concerns from other research about SARS-CoV-2 lineages carrying L452R.

The direct protection provided by COVID-19 vaccines against hospitalization and death in particular has been highlighted as among the most important vaccine efficacy endpoints (17). Importantly, while hospitalizations following SARS-CoV-2 infection in vaccinated persons were rare in this study, we find that the association of E484K with infection in vaccinated persons persisted even if we restricted the outcome only to persons who were hospitalized within 28 days of infection. Given the small number of hospitalizations following infection in vaccinated persons observed in this study additional studies focused on severe illness are needed.

We also estimated separately the association of each substitution with infection in vaccinated persons with each of the three vaccines in use in Maryland. While there were differences in these crude associations between vaccines, there was substantial overlap in the confidence intervals for these estimates. The data here are not sufficient to determine whether some vaccines are more effective against viruses carrying the E484K or L452R substitutions than others.

Finally, it is important to note that during the period of this study, the rate of confirmed COVID-19 cases in Maryland fell 10-fold (18). At the same time, the fully vaccinated increased from fewer than 1,000 to approximately 2.8 million in a population of approximately 6 million (22). In this study, in which all members of the study cohort were infected with COVID-19, more than 97% of persons were not fully vaccinated against COVID-19 at the time of their infection. Nothing here should be interpreted to suggest that mass vaccination campaigns will not be effective for reducing the incidence of COVID-19.

This analysis has a number of limitations. 1) This study, which included only vaccinated persons, does not directly estimate vaccine effectiveness; that would require a comparison of the rate of infection between vaccinated and unvaccinated people. 2) The population of people infected with SARS-CoV-2 in this study may not be representative of the population of people infected with SARS-CoV-2 in Maryland during the same period. Only some laboratories either conduct sequencing or make specimens available to the Maryland Department of Health laboratory for sequencing; populations tested by other laboratories would not be represented in this analysis. Further, infections in vaccinated persons were preferentially selected for sequencing by Maryland’s public health lab. 3) This study examined two specific SARS-CoV-2 spike protein mutations. SARS-CoV-2 mutations are not randomly assorted; if there is some other spike protein mutation that commonly co-occurs with E484K or L452R in Maryland, that mutation could actually be responsible for the associations observed here. 4) Data on patients’ underlying health conditions were not available. 5) In analyses of hospitalization, the outcome was hospitalization within 28 days of infection. The cause of hospitalization was not available; some hospitalizations may not have been caused by SARS-CoV-2 infection.

This analysis is the first to examine the association of the E484K and L452R substitutions with SARS-CoV-2 infection and subsequent hospitalization in persons who received a COVID-19 vaccine. It provides evidence current vaccines may be less effective at preventing both infection and hospitalization from SARS-CoV-2 viruses that carry the E484K substitution. These findings underscore the importance of conducting SARS-CoV-2 genomic surveillance and integrating those data with other surveillance data to quickly identify mutations that may be eroding the effectiveness of vaccines.

## Data Availability

Data are the property of the U.S. state of Maryland and are collected for public health surveillance purposes. They are not available to the public.

## Acknowledgments

We wish to acknowledge the following groups that submitted specimens to GISAID:

- Dakota Howard, Dhwani Batra, Peter W. Cook, Kara Moser, Adrian Paskey, Jason Caravas, Benjamin Rambo-Martin, Shatavia Morrison, Christopher Gulvick, Scott Sammons, Yvette Unoarumhi, Darlene Wagner, Matthew Schmerer,Matthew Tugwell, Lauren Moon, Clinton R. Paden, Duncan MacCannell. Centers for Disease Control and Prevention Division of Viral Diseases, Pathogen Discovery.
- Tallon, Luke J; Sadzewicz, Lisa D; Humphrys, Mike; Ott, Sandra; Roussey, Holly; Mehta, Aditya; Vavikolanu, Kranthi; Fraser, Claire M; Ravel, Jacques. Maryland Genomics, Institute for Genome Sciences, University of Maryland School of Medicine
- C. Paul Morris, Chun Huai Luo, Adannaya Amadi, Matthew Schwartz, Nicholas Gallagher, Heba H. Mostafa. Johns Hopkins Hospital Department of Pathology.
- Maryland Department of Health Laboratories Administration.

This research was supported in part by the National Institutes of Health Intramural Research Program.

The findings and conclusions in this report are those of the authors and do not necessarily represent the official position of the Centers for Disease Control and Prevention. Use of trade names and commercial sources is for identification only and does not imply endorsement by the Centers for Disease Control and Prevention, the Public Health Service, or the U.S. Department of Health and Human Services.

